# Clinical Outcome of Neurological patients with COVID-19: the impact of Healthcare organization improvement between waves

**DOI:** 10.1101/2021.11.19.21266606

**Authors:** Viviana Cristillo, Andrea Pilotto, Alberto Benussi, Ilenia Libri, Marcello Giunta, Andrea Morotti, Stefano Gipponi, Martina Locatelli, Stefano Cotti Piccinelli, Valentina Mazzoleni, Francesca Schiano di Cola, Stefano Masciocchi, Debora Pezzini, Andrea Scalvini, Enrico Premi, Elisabetta Cottini, Massimo Gamba, Mauro Magoni, Marco Maria Fontanella, Alessandro Padovani

**Affiliations:** Department of Clinical and Experimental Sciences, Neurology Unit, University of Brescia, Italy; Stroke Unit, Azienda Socio Sanitaria Territoriale Spedali Civili, Spedali Civili Hospital, Brescia, Italy; Neurosurgery Unit, Department of Medical and Surgical Specialties, Radiological Sciences and Public Health, University of Brescia, Spedali Civili di Brescia, Brescia, Italy

**Keywords:** COVID-19, neurological disease, outcomes, steroid therapy, mortality

## Abstract

**Objective:** The aim of this study is to evaluate the differences of clinical presentations and the impact of healthcare organization on outcomes of neurological COVID-19 patients admitted during the first and second pandemic waves.

**Methods:** In this single center cohort study, we included all patients with SARS-CoV-2 infection admitted to a Neuro-COVID Unit. Demographic, clinical and laboratory data were compared between patients admitted during the first and second waves of COVID-19 pandemic.

**Results:** 223 patients were included, of whom 112 and 111 hospitalized during the first and second pandemic waves, respectively. Patients admitted during the second wave were younger and exhibited pulmonary COVID-19 severity, resulting in less oxygen support (n=41, 36.9% *vs* n=79, 70.5%, p<0.001) and lower mortality rates (14.4% *vs* 31.3%, p=0.004). The different healthcare strategies and early steroid treatment emerged as significant predictors of mortality independently from age, premorbid conditions and COVID-19 severity in cox regression analyses.

**Conclusions:** Differences in healthcare strategies during the second phase of COVID-19 pandemic probably explain the differences in clinical outcomes independently of disease severity, underlying the importance of standardized early management of neurological patients with SARS-CoV-2 infection.

## Introduction

Coronavirus disease-19 (COVID-19), associated with the severe acute respiratory syndrome Coronavirus 2 (SARS-CoV-2), has become a global pandemic, giving rise to a serious health burden globally. Many countries worldwide experienced a two-wave pattern of COVID-19 spreading during the pandemic, with a first wave during spring 2020[1] followed by the second wave starting in late summer 2020 persisting until the spring of 2021. Neurological symptoms and syndromes concomitant SARS coV-2 infection have been associated with increased risk of mortality and poor outcome in independent case series[1-4]. Recent data from general COVID Units suggested that the patients hospitalized during the first and second waves of COVID-19 pandemic differed for age range, severity of the disease, COVID-19 treatment strategies adopted and outcomes[5] but no data specifically evaluating neurological patients are still available.

In this work, we aimed to evaluate the impact of different healthcare strategies on final outcomes. by compare clinical and laboratory characteristics of hospitalized COVID -19 neurological patients during the first and second waves of pandemic in a single territory hub for Neuro-COVID patients.

## Methods

### Study design and participants

This cohort study included adult inpatients (≥18 years old) with SARS-CoV2 infection admitted at the Neuro-COVID Unit of the ASST Spedali Civili Hospital, Brescia, for neurological diseases from February 21 to June 5, 2020 (first wave) and from November 1 to April 30, 2021(second wave). SARS-CoV-2 infection was confirmed by RT-PCR in nasopharyngeal/oropharyngeal swabs or bronchoalveolar lavage. The study received approval from local ethics committee of the ASST Spedali Civili Hospital, Brescia (NP 4067, approved 08.05. 2020). Premorbid conditions were recorded at admission using the Cumulative Illness rating scale (CIRS) and the pre-morbid modified Rankin scale (mRS). Hospitalization data included the severity of COVID-19 disease, expressed by the Brescia-COVID Respiratory Severity Scale (BCRSS)[6] and the quick Sequential Organ Failure Assessment (qSOFA) score.

Steroid treatment with methylprednisolone 1 g/day for five days was defined as high-dose treatment (HDS), whereas dexamethasone 6 mg/day was defined as standard-dose treatment (SDT). During the second wave, the health organisation system changed adopting a) different referral system from family doctors for patients at higher risk of deterioration b) standardisation of patients profiling using neurological, comorbid and frailty measures c) standardisation of management of neurological patients in COVID-19 including specific internal guidelines for stroke, encephalitis, delirium, seizures, headache d) multidisciplinary team of clinicians including neurologist, internal medicine and infectious disease specialists in the unit e) early use of steroid and heparin according to updated COVID-19 guidelines[7] f) larger use of non-invasive ventilation in non-ICU units g) early screening for ICU need.

### Statistical analysis

Continuous and categorical variables are reported as mean values ± standard deviation and n (%) respectively. Differences between patients during the two waves were compared by t-test or Fisher’s exact test where appropriate. Linear regression models adjusted for the effect of age, COVID-19 severity, comorbidities and baseline mRS evaluated the impact on different waves on mRS at discharge. Cox regression model based on previous findings[1] was implemented in order to investigate the combined effect of predictors of mortality, namely age, qSOFA scores, BCRSS, platelet count, first vs second wave, steroid treatment, time from symptoms onset to admission. A two-sided p-value<0.05 was considered significant; data analyses were carried out using SPSS software (version 21.0).

## Results

Two hundred twenty-three COVID-19 patients were hospitalized in the Neuro-COVID Unit of the ASST Spedali Civili di Brescia Hospital, of whom 112 admitted from February 21 to June 5, 2020 and 111 hospitalized from November 1 to April 30, 2021.

Demographic, clinical and laboratory characteristics of included patients are reported in Table 1. Patients admitted during the second wave were younger (years 62.9+18.9 *vs* 72.6±12.1, p<0.001), exhibited a lower comorbidity severity index (1.21±0.2 *vs*. 1.28±0.2, p=0.026), less severe pulmonary disease, expressed by lower qSOFA score (0.48±0.7 *vs* 0.87±0.7, p<0.001) and lower BCRSS (0.50±0.8 vs. 1.24±0.97, p<0.001) at admission compared to patients hospitalized during the first outbreak.

**Table 1.**
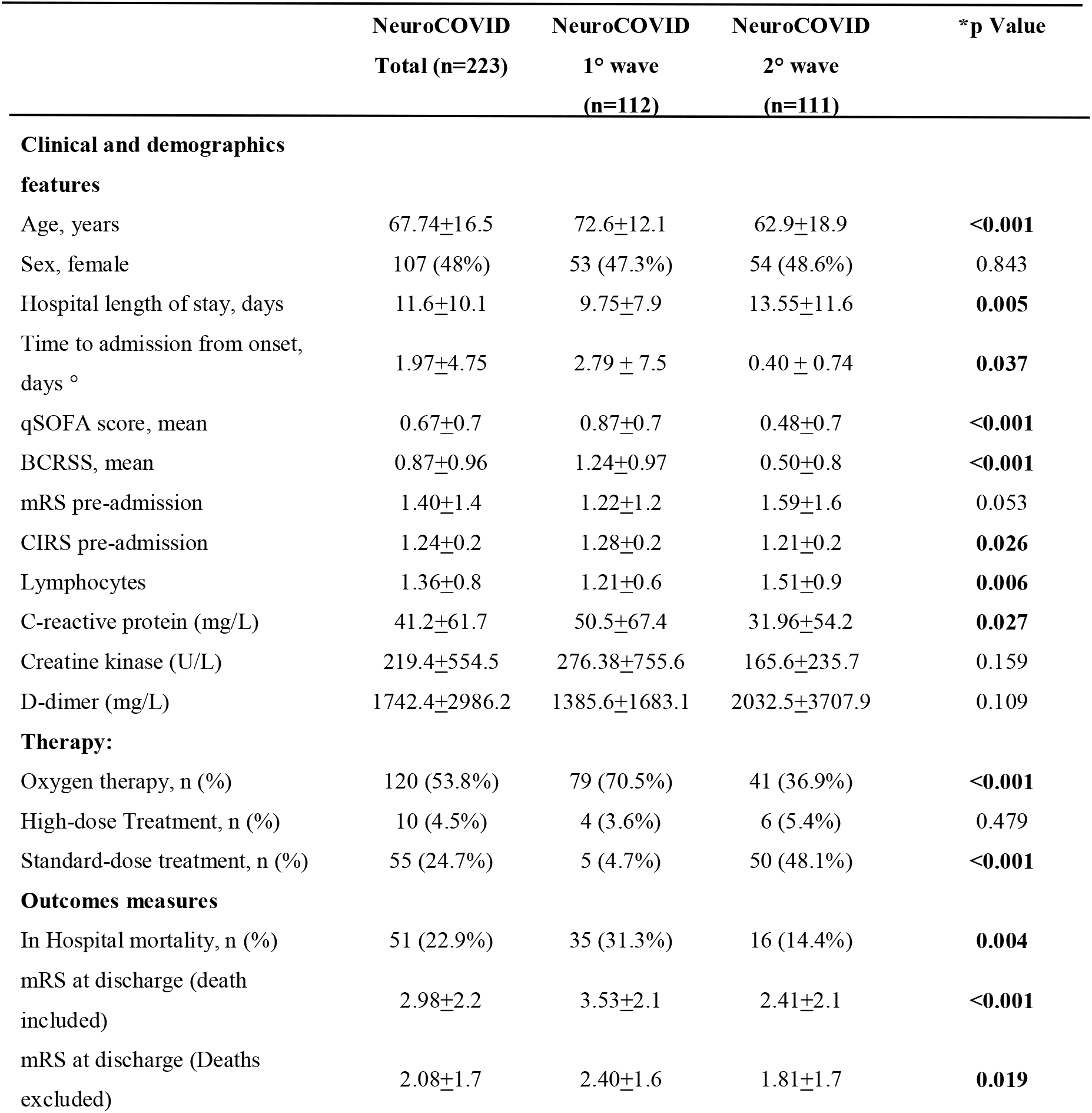
Demographic, clinical, laboratory characteristics patients according to first and second pandemic waves. \**p* values were calculated by t-test or Fisher’s exact test, as appropriate; ° patients with BCRSS ≥ 2; Abbreviations: BCRSS, Brescia-COVID Respiratory Severity Scale; CIRS, Cumulative Illness Rating Scale; GBS, Guillain-Barrè syndrome; mRS, modified Rankin scale; qSOFA, quick sequential organ failure assessment.

COVID-19 patients admitted during the first wave showed higher blood inflammatory parameters, chest X-Ray scores (Table 1) and use of high flow oxygenation (n=79, 70.5%, n=41, 36.9% *vs* p<0.001) compared to patients admitted during the second outbreak.

The specific neurological diagnosis exhibited a different distribution within the two time-periods (Figure 1) characterized by a significant reduction in stroke rates during the second wave (n=54, 48.2% vs n=25, 22.5%, p<0.001). Patients with moderate to severe respiratory disease exhibited a shorter time from symptom onset to hospitalization in the second wave compared to the first pandemic phases (days 0.40 ± 0.74 vs 2.79 ± 7.5, p=0.037).During the second wave, patients showed lower mortality rates after adjusting for age and COVID-19 severity (n=32, 38.6% vs n=19, 17.1%, p=0.009), and better clinical outcomes adjusting for baseline status (mRS at discharge 2.40 ± 1.6 vs 1.81 ± 1.7, p=0.019).

**Figure 1:**
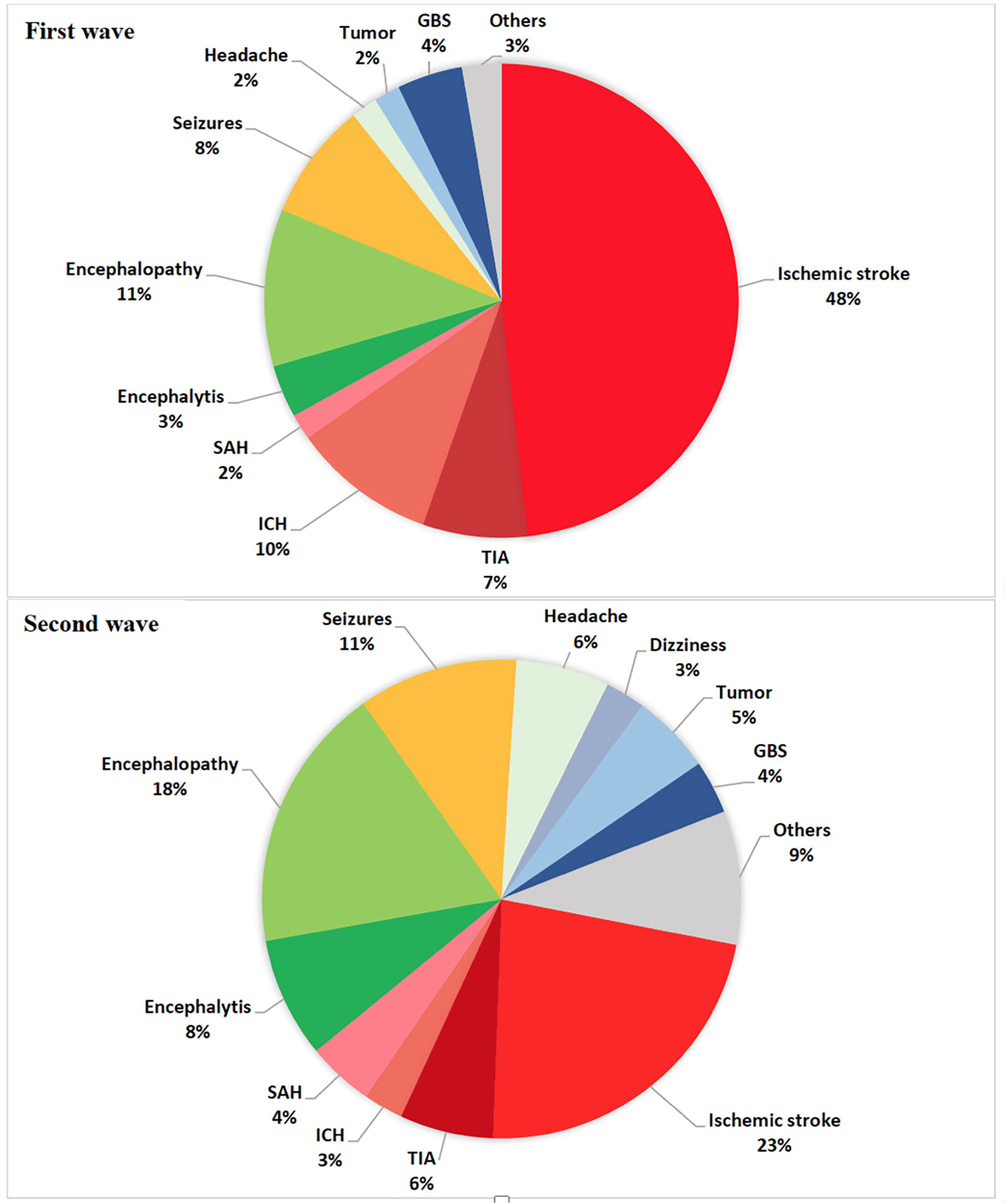
Neurological diagnosis distribution during the first and second pandemic waves. Abbreviations: GBS, Guillain-Barrè syndrome; ICH, Intracerebral haemorrhage, SAH, Subarachnoid haemorrhage; TIA: transient ischaemic attack.

Cox regression model identified age (p=0.001), COVID-19 severity (i.e BCRSS, p<0.001), pre-morbid comorbidity (i.e. CIRS, p=0.028) and the different time-period (i.e. waves, p=0.012) as independent significant predictors of mortality in hospitalized patients. Standard steroid treatment was adopted in 4.7 % and 48.1% of patients admitted during the first and second waves, respectively (p<0.001). Specific COX-regression analyses revealed steroid treatment as independent predictor of survival (ExpB 2.084, IC 1.072-4.050, p=0.007) after adjusting for age, BCRSS, CIRS and time period-in the global sample (Supplementary figure 1).

## Discussion

The study showed that patients admitted during the second wave of COVID-19 pandemic were younger, exhibited lower pulmonary severity, a different distribution of neurological diagnosis, lower mortality rates and better neurological outcomes compared to patients admitted during the first wave. The different healthcare strategies adopted during the two phases of the pandemic and the modulation with steroid treatments appeared to be independent predictors of mortality in addition to age, pre-morbid conditions and COVID-19 severity in the cohort.

This study included 223 consecutive COVID-19 patients hospitalized for neurological disorders admitted during two different peaks of pandemic in Italy in a tertiary hub for Neuro-COVID patient. During the first phases of the pandemic, we observed a higher prevalence of cerebrovascular diseases, representing more than half of patients evaluated in the emergency room[8]. During the second phase of the pandemic, conversely, we observed slightly higher prevalence of patients hospitalized for encephalopathies or headache, whereas cerebrovascular events decrease to about a third of admitted patients. These differences might be due to younger age, lower comorbidity status and lower severity of COVID-19 observed during the second wave, as severe SARS-Cov-2 infection with prominent systemic response have been claimed to be associated with increased risk of stroke[2-4]. The Cox regression model identified the severity of pulmonary disease, age and pre-morbid conditions as the most important predictors of mortality, with a strong difference in mortality rates between first and second waves (38% vs 17%). On one hand, the general decrease of severity of COVID-19 disease during the second wave-reported in Europe and United Stated – might largely explain the reduction of mortality and medical complications observed[9-12]. On the other, the management of COVID-19 patients consistently improved during the second pandemic wave both at primary care and at hospital level[13]. The total number of COVID-19 dedicated Units and beds were increased along with both the non-invasive ventilation and patients were hospitalized earlier and with milder symptoms[10]. Second, the lessons learned from the first phases of pandemic allowed the development of standardized procedures and guidelines for the management of patients with moderate and severe COVID-19 disease, thus strongly improving the care of patients since the early stages. Third, the primary care doctors were directly involved in the initial management of patients with COVID-19 infection thus increasing the referral to specific COVID-19 Units when needed[11]. Dexamethasone and remdesivir substituted hydroxychloroquine and lopinavir, and anticoagulation therapy was promptly administrated since the first days of admission[14]. Indeed, the time interval since onset appeared to improve survival and outcome independently from the severity of disease. Furthermore, the known increased inflammatory response to the viral infection might be an important modulator of incidence and severity of related CNS disorders[2-4,15] in addition to systemic complications. This fits with the results of our study, indicating immunomodulatory treatment as independent predictor of mortality -in addition to the time of hospitalization and severity of the disease.

We acknowledge that this work entails some limitations, as this is a monocentric study with a relative small sample size and we could not exclude that some patients with COVID-19 disease and neurological symptoms or syndromes did escape the referral, especially for mild cases not requiring hospitalization.

Nevertheless, this is the first study evaluating the differences between neurological COVID-19 patients during the two pandemic waves. Findings showed that different management strategies adopted and the lessons learned by health workers from the first pandemic phases largely explain the improvement in final outcomes observed independently from the reduction of severity of SARS-CoV-2 infection. Larger on-going multicenter studies are warranted to confirm and extend these findings in order to understand the future global impact of healthcare system organization, immunomodulator treatments and the large use of vaccination on the outcome of neurological patients with COVID-19 disease.

## Data Availability

All data produced in the present work are contained in the manuscript

## Notes

**Financial disclosure and conflict of interest regarding the research related to the manuscript:** All authors have no conflict of interest regarding the research related to the manuscript. The full financial disclosure for the past year (for all authors) is documented at the end of the manuscript.

**Study funding:** The study was not financial supported

### Competing Interest Statement

The authors have declared no competing interest.

### Funding Statement

This study did not receive any funding

### Author Declarations

Ethics committee of University of Brescia gave ethical approval for this work

