## Supplementary figure 1; Supplementary table 1 for "Clinical Outcome of Neurological patients with COVID-19: the impact of Healthcare organization improvement between waves"

**Supplementary figure 1** Cox Regression model evaluating the impact of hospitalization period (i.e first vs second pandemic wave, Panel A) and Steroid Treatment (Panel B). Data are corrected for the effect of age, sex, premorbid comorbidity index, COVID-19 disease severity.

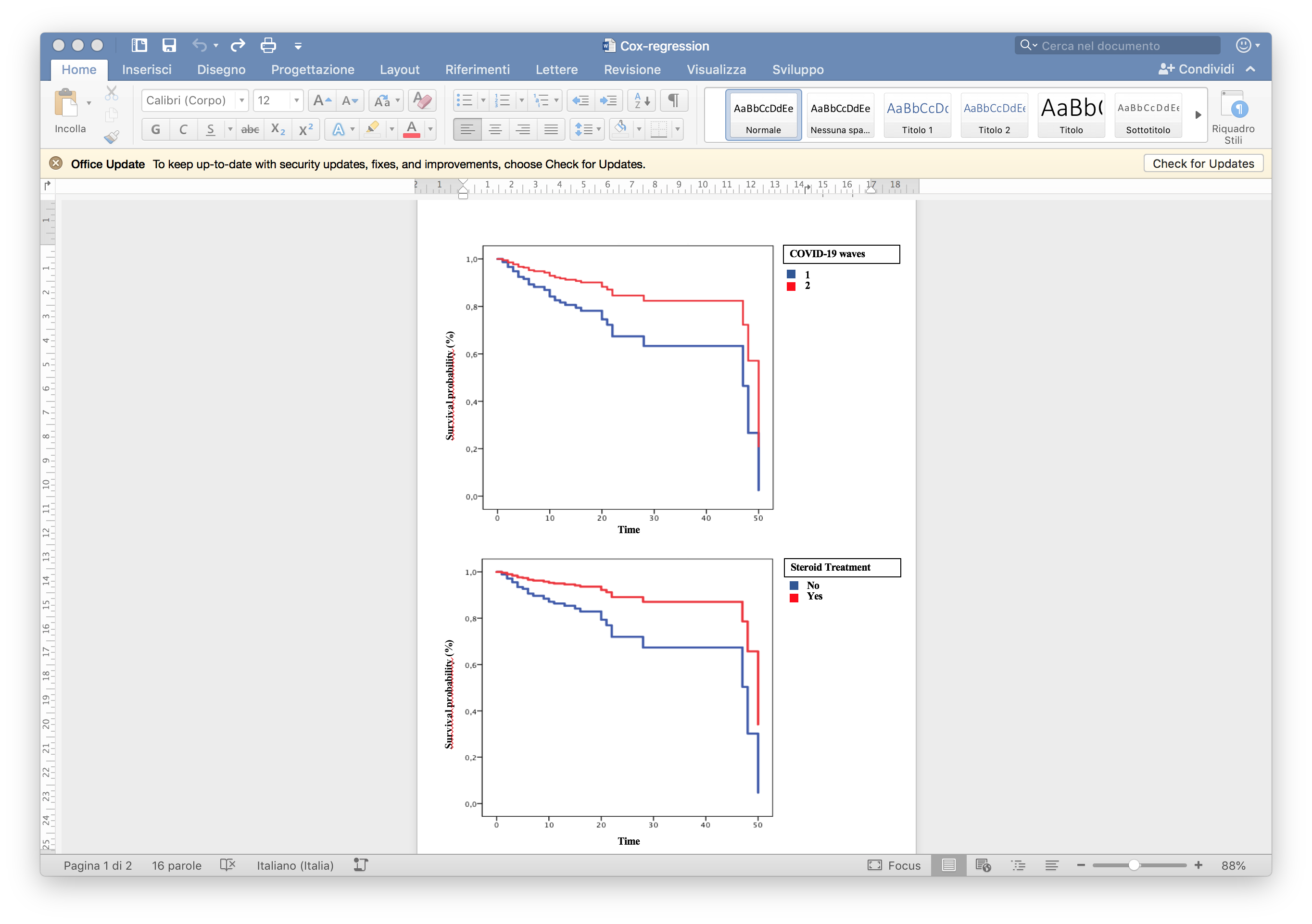

**Supplementrary Table 1** Neurological diagnosis distribution during the first and second pandemic waves. Abbreviations: GBS, Guillain-Barrè syndrome; ICH, Intracerebral haemorrhage, SAH, Subarachnoid haemorrhage; TIA: transient ischemic attack.

|  | **NeuroCOVID** | **NeuroCOVID** | **NeuroCOVID** | ***p Value** |
| --- | --- | --- | --- | --- |
|  | **Total (n=223)** | **1° wave** | **2° wave** |  |
|  |  | **(n=112)** | **(n=111)** |  |
| **Admitting diagnosis** |  |  |  |  |
| Stroke | 79 (35.4%) | 54 (48.2%) | 25 (22.5%) | **<0.001** |
| ICH | 14 (6.3%) | 11 (9.9%) | 3 (2.7%) | 0.050 |
| SAH | 7 (3.1%) | 2 (1.8%) | 5 (4.5%) | 0.280 |
| TIA | 15 (6.8%) | 8 (7.2%) | 7 (6.3%) | 0.789 |
| Seizures | 20 (9.0%) | 9 (8.0%) | 12 (10.8%) | 0.501 |
| encephalitis | 13 (5.8%) | 4 (3.6%) | 9 (8.1%) | 0.166 |
| Encephalopathy | 14 (6.3%) | 12 (10.7%) | 20 (18.0%) | 0.050 |
| Tumor | 8 (3.6%) | 2 (1.8%) | 6 (5.4%) | 0.171 |
| Headache | 9 (4.0%) | 2 (1.8%) | 7 (6.3%) | 0.101 |
| Dizziness | 3 (1.3%) | 0 | 3 (2.7%) | 0.122 |
| GBS | 9 (4.0%) | 5 (4.5%) | 4 (3.6%) | 0.744 |
| Others | 13 (5.8%) | 3 (2.7%) | 10 (9.0%) | 0.050 |
